# Developing a phenotype risk score for *TTR* V142I to capture undiagnosed variant transthyretin amyloidosis in health systems

**DOI:** 10.64898/2026.01.05.26343489

**Authors:** Deepa Sarkar, Kathleen D. Ferar, Mohammad Ghouse Syed, Lisa A. Bastarache, Eimear E. Kenny, Noura S. Abul-Husn, Vikas Pejaver, Amy R. Kontorovich

## Abstract

**Background:** Phenotype Risk Scores (PheRS) leverage electronic health record (EHR) data to identify individuals at risk for Mendelian disorders, but their performance remains untested for diseases with common and/or non-specific features such as variant transthyretin amyloidosis (ATTRv), often presenting with heart failure (HF), atrial fibrillation, polyneuropathy, and other prevalent diagnoses. We optimized a PheRS for the most common form of ATTRv by integrating genomic and clinical data in Mount Sinai’s Bio*Me* biobank, focusing on expert-driven phenotype definitions for the *TTR* variant p.Val142Ile (V142I), which is prevalent in African American (AA) populations (4%).

**Methods:** We developed and evaluated a customized PheRS for ATTRv that incorporated 21 expert-curated phenotypic features including 292 ICD-9 and ICD-10 diagnosis codes on a biobank cohort of V142I+ cases (n=383) and controls without any pathogenic/likely pathogenic *TTR* variants (n=30,642). We compared its performance with the standard automated PheRS approach using different metrics. To account for age-dependent penetrance and high lifelong risk of HF, we further tested the customized PheRS for V142I in a subset of individuals of age ≥ 60 with self-reported Black or AA race/ethnicity and at least one occurrence of HF in their EHRs.

**Results:** The expert-curated PheRS outperformed the standard PheRS as measured by improved precision-at-*k* (0.05 vs. 0.00; *k*=100), a demonstrably, clinically relevant metric. In the subcohort enriched for anticipated penetrance (older, Black/AA HF patients), the expert-curated PheRS identified more V142I+ individuals (6.0%) among the top 100-scoring individuals than a strategy that randomly sampled from the population (3.6%).

**Conclusion:** This work demonstrates that standard PheRS methods are insufficient for common, adult-onset cardiovascular genetic diseases such as V142I-related ATTRv, but when redesigned with disease biology, ancestry, age, and clinical context in mind, PheRS become clinically actionable tools for precision cardiology.

## INTRODUCTION

Risk scores derived from electronic health record (EHR) phenotyping have emerged as a promising class of data-driven methods for the identification of individuals at increased risk for Mendelian diseases.^1–4^ These scores quantify how closely a patient’s clinical presentation reflected through diagnosis codes in their EHRs matches the known phenotype of a specific genetic disorder. The fundamental idea behind the Phenotype Risk Score (PheRS), one of the earliest approaches, is that many of the clinical manifestations of rare diseases are, by definition, uncommon in the general patient population.^1^ By assigning higher weights to these rarer features, PheRS capture the likelihood that a patient’s clinical profile is indicative of a particular Mendelian condition. Since their original publication, the PheRS calculation approaches have been modified and adapted for various diseases and applications in clinical and research settings.^1,5–10^ They are being used to: 1) prioritize individuals for genetic testing where traditional risk factors or family history may miss at-risk individuals; 2) identify potentially undiagnosed cases within large healthcare systems by scanning for phenotypic signatures; and 3) construct cohorts for the evaluation of the pathogenicity of variants in genotype-first studies.

Despite promising advances in the use of PheRS, their effectiveness for genetic conditions presenting with common or non-specific symptoms remains uncertain, such as the majority of monogenic cardiovascular diseases, including those on the American College of Medical Genetics’ (ACMG) list of secondary finding genes. The assumption of the rarity of monogenic disorder-related clinical features that underlies PheRS may not hold and such overlap with common conditions can dilute the discriminatory power of traditional PheRS methods. Furthermore, the clinical utility of these methods in prioritizing individuals for genetic testing remains understudied, especially for conditions caused by variants that may be specific to certain ancestries and/or incompletely penetrant.

Hereditary (variant) transthyretin amyloidosis (ATTRv) is an exemplar disorder of a cardiovascular genetic condition with common and non-specific symptoms. ATTRv is an adult-onset progressive disorder due to pathogenic variants in the *TTR* gene that may manifest with multiple features that are relatively common in an adult health system population, including cardiomyopathy, heart failure (HF), peripheral neuropathy, and carpal tunnel syndrome. More than 140 disease-causing *TTR* variants have been identified to date, and highly specific genotype-phenotype associations are well described across variants.^11^ The most common pathogenic ATTRv variant globally, p.Val142Ile (V142I), is present in up to 4% of Black and/or African American individuals and up to 1% of Hispanic or Latino individuals in the United States.^12,13^ V142I manifests disease predominantly in adults age 60 or older,^14–16^ and depending on age, its penetrance is estimated to be 37.4-38.8%,^14,17^ which may be lower than other *TTR* variants, particularly the European-predominant p.Val50Met and p.Thr80Ala alleles. In general, ATTRv is one of the most striking examples of age-related penetrance among monogenic conditions.

To move PheRS from promise to practice in adult-onset monogenic diseases with common clinical features, we evaluate ATTRv as a test case and ask: Which clinical features, evaluation metrics, and deployment contexts make PheRS clinically useful? We contrast a standard phecode-based PheRS with an expert-curated ICD feature set, adopt precision-at-*k* as a case-finding endpoint aligned with downstream testing, and evaluate performance for two other cardiovascular genetic conditions, Marfan and Long QT syndromes. We find that disease-informed curation and context-aware deployment improve actionable precision relative to standard approaches, and we outline a practical blueprint for implementing PheRS to identify undiagnosed patients, particularly in populations bearing the greatest burden of missed diagnoses.

## METHODS

### Integration of electronic health record and genomic data

The Bio*Me* Biobank is an electronic health record (EHR)-linked biobank of over 60,000 participants recruited predominantly through ambulatory care practices across the Mount Sinai Health System in New York, New York. All Bio*Me* participants provided written informed consent , with approval by the Icahn School of Medicine at Mount Sinai (ISSMS) Institutional Review Board (IRB 07–0529). Under the current ISSMS IRB-approved study (STUDY-18-00223), EHR data for all individuals within the Bio*Me* biobank was obtained from the Mount Sinai Data Warehouse (MSDW) via an honest broker mechanism. For this analysis, demographic variables (year of birth, sex, race, and ethnicity), diagnosis information (ICD-9 and ICD-10 codes and descriptions) and encounter variables (date and time of ICD codes), were used. *TTR* variant identification from sequencing/genotyping data and linkage to EHR data was performed as before.^14^

### Custom phenotypic risk score (PheRS) approach

ICD-9 and ICD-10 codes relevant to ATTRv were identified from the literature^13,18,19^ and known disease presentation patterns. These were then programmatically expanded to include possibly relevant codes from within the ICD-9 and ICD-10 code hierarchies. Finally, clinical experts within the author group iteratively removed irrelevant ICD codes and grouped the remaining codes into clinical features or phenotypes. This resulted in 292 ICD codes being mapped to 21 phenotypes (Table S1). A PheRS calculator was implemented in Python following Bastarache *et al*., assigning weights equal to the log-transformed inverse frequency of each phenotype in a training population (as described below), thus, assigning higher weights to rarer phenotypes. For each individual, the presence or absence of a phenotype was determined based on the occurrence of at least one of the corresponding ICD codes in their health record, and the total PheRS was the sum of the weights for all observed phenotypes.

### PheRS and algorithmic variations

The original PheRS approach was applied as a comparator through the “phers” R library. This approach differs mainly in its automated definition of phenotypes using phecodes, which aggregate ICD codes in a disease-agnostic manner. Only phecodes mapped to human phenotype ontology (HPO) terms for the target Mendelian disease (via OMIM) are used for PheRS computation. For ATTRv, the identifier that was input was 105210. The library also allows for the calculation of alternate versions of PheRS, which are referred to here as algorithmic variations. Among these, two were considered for this study: (1) PheRS with negative weights, which penalizes the absence of disease-related phecodes rather than assigning zero weights to them; and (2) PheRS with precomputed weights from Vanderbilt University’s BioVU biobank, testing generalizability across health systems.

### Cohorts for PheRS weight derivation and calculation

Because specific genotype-phenotype associations are well-described in ATTRv^20^ and the vast majority of carriers in our health system harbor the c.424G>A, p.(Val142Ile) variant (V142I) in the *TTR* gene, we restricted our analyses to this variant. The case group included individuals harboring this variant, as observed in genotype array or exome sequencing data. The control group included any individual without a likely pathogenic or pathogenic variant in *TTR* as recorded in ClinVar (release 20240611); this group was defined solely based on exome sequencing data to ensure the actual absence of these variants.

A cohort was constructed for the sole purpose of calculating PheRS weights, i.e., a “training” cohort. All individuals in the Bio*Me* biobank who were not assigned to the case and control groups were assigned to this “training” cohort, regardless of their genotype status. Since this cohort was orders of magnitude smaller than the actual patient population at Mount Sinai and not necessarily representative of the general patient population, an alternative “training” cohort was constructed for additional analyses. For this, ICD code data for 4,373,534 patients from the Mount Sinai Data Warehouse (MSDW) were directly extracted via the AI Ready Mount Sinai (AIR.MS) platform.^21^

### Evaluation metrics

To assess the ability of PheRS to distinguish between variant-positive and variant-negative individuals, each approach was evaluated using a suite of performance metrics. *P*-values to test for significant differences between the case and control PheRS distributions were calculated using the non-parametric Wilcoxon rank sum test, as is typical in the PheRS literature.^11,22^Additionally, the area under the receiver operating characteristic curve (AUC), a standard metric in machine learning and risk score applications, was calculated for each PheRS approach to measure its discriminative ability. Finally, motivated by the objective of prioritizing clinically actionable patients from a cohort, the positive predictive value or precision was calculated for the top *k*-scoring individuals in our cohorts, i.e., precision-at-*k*, with varying values of *k*. The Wilcoxon rank sum test and AUC evaluate whether the distribution of values for affected individuals is significantly different from that of unaffected individuals. Precision-at-*k* assesses the relevance of individuals with very high scores, simulating a scenario where the top *k*-scoring individuals are selected for screening in a prospective evaluation. For this study, *k* was initially chosen to be 100, further experiments were carried out at multiple values for *k* ranging from 10 to the entire cohort size to identify the optimal range of values.

### Extension to other cardiovascular diseases

We also tested PheRS applications in our health system for two other monogenic cardiovascular diseases: Marfan syndrome and Long QT syndrome. These were selected because they are expected to have relatively high penetrance, strong clinical significance and are genetically represented on the ACMG secondary findings list. For each disease, cases were defined as those individuals harboring a likely pathogenic or pathogenic variant in a gene associated with the disease, as classified previously by a “badge” laboratory in ClinVar.^23^ The genes considered were *FBN1* for Marfan syndrome and *KCNQ1*, *KCNH2* and *SCN5A* for Long QT syndrome. Controls were defined in a manner similar to that for ATTRv. A custom PheRS was also developed similarly to ATTRv by mapping ICD codes to clinical features or phenotypes (Tables S2 and S3). Any ICD codes that mapped to an actual diagnosis of the disease were excluded from PheRS calculation to avoid information leakage, e.g., Q87.4 and its descendant codes in ICD-10 for Marfan syndrome. The original PheRS approach was run by using 154700 and 192500 as the OMIM identifiers for Marfan and Long QT syndromes respectively. While there were multiple identifiers associated with Long QT syndrome in OMIM, each corresponding to a type of the disorder based on the underlying gene, a single identifier (corresponding to LQT1) was chosen as no differences in PheRS performance were observed between OMIM identifiers (data not shown).

## RESULTS

### Baseline demographic characteristics of cohorts

We first compared demographic characteristics between cases and controls for each of the three cohorts (Table 1). No meaningful differences were observed for age, race/ethnicity and sex, except for the unsurprising overrepresentation of Black or African American individuals among V142I+ cases. Due to our focus on ATTRv, we further tested for associations between V142I+ cases and controls with other cardiovascular disease-related comorbidities for the full cohort as well as Black/African American individuals (Table S4). In both cohorts, the occurrence of these comorbidities were generally balanced across cases and controls.

**Table 1.** Summary of cohort sizes for all three diseases.

|  | ATTRv |  | Marfan syndrome |  | Long QT syndrome |  |
| --- | --- | --- | --- | --- | --- | --- |
|  | Cases<br>(n=393) | Controls<br>(n=30396) | Cases<br>(n=29) | Controls<br>(n=30766) | Cases<br>(n=42) | Controls<br>(n=30752) |
| **Study Cohort,**<br>**n (%)** |  |  |  |  |  |  |
| ICD codes<br>available | 383 (97.5) | 29266 (96.3) | 28 (96.6) | 29625 (96.3) | 42(100) | 29611(96.3) |
| **Patient Characteristics** |  |  |  |  |  |  |
| Mean age (SD) | 65.3 (16.7) | 64.6 (17.4) | 61.2 (17.1) | 64.6 (17.4) | 64.2 (15.7) | 64.8 (17.4) |
| **EHR linked**<br>**Race/ethnicity**<br>**n (%)** |  |  |  |  |  |  |
| BLACK OR<br>AFRICAN<br>AMERICAN | 215 (56.2) | 6482 (22.2) | 9 (32.1) | 6690 (22.6) | 8 (19) | 6691 (22.6) |
| HISPANIC<br>OR<br>LATINO | 66 (17.2) | 5148 (17.6) | 5 (17.9) | 5206 (17.6) | 9 (21.4) | 5202 (17.6) |
| WHITE | 3 (0.8) | 8557 (29.2) | 6 (21.4) | 8558 (28.9) | 10 (23.8) | 8554 (28.9) |
| OTHERS | 99 (25.9) | 9079 (31) | 8 (28.6) | 9171 (31) | 15 (35.7) | 9164 (30.9) |
| **Sex**<br>**n (%)** |  |  |  |  |  |  |
| Female | 235 (61.5) | 17341 (59.3) | 21 (75.0) | 17563 (59.3) | 25 (59.5) | 17559 (59.3) |
| Male | 148 (38.5) | 11911 (40.69) | 7 (25) | 12048 (40.7) | 17 (40.5) | 12038 (40.7) |
| Others | 0 (0) | 14 (0.1) | 0 (0) | 14 (0.1) | 0 (0) | 14 (0.1) |

### Custom PheRS approach outperformed standard PheRS approaches in terms of clinical utility

We first developed a custom PheRS approach for ATTRv that uses the same inverse prevalence-based weighting scheme as proposed by Bastarache et al. but does not rely on phecodes or other mapped vocabularies.^11,24^ Instead, our approach directly applies this scheme to groups of ICD-9 and ICD-10 codes constructed based on expert knowledge to reflect distinct and specific phenotypic features of ATTRv (Table S1). We compared this custom PheRS to the original PheRS approach developed by Bastarache *et al.* and its algorithmic variations.

PheRS are typically assessed in terms of their ability to distinguish between cases and controls through a Wilcoxon rank sum test. Using this metric, the custom and original PheRS both resulted in significantly higher scores for cases than for controls: 1.214 vs. 0.997 and 1.408 vs. 0.851, respectively (Table 2). This was true for the original PheRS when negative weights for the absence of certain phecodes were allowed (0.963 vs. 0.454) and when precomputed weights from a different biobank were used (1.810 vs. 1.268). However, such statistical tests are prone to Type I errors on highly imbalanced case-control cohorts, such as those in this study, resulting in significant *P*-values even when case and control score distributions greatly overlap (Figure 1). Therefore, we also compared AUCs, a metric that is more robust to case-control imbalance. All four approaches yielded AUC values marginally greater than 0.5, with the original PheRS and its algorithmic variations modestly outperforming the custom PheRS.

**Figure 1.**
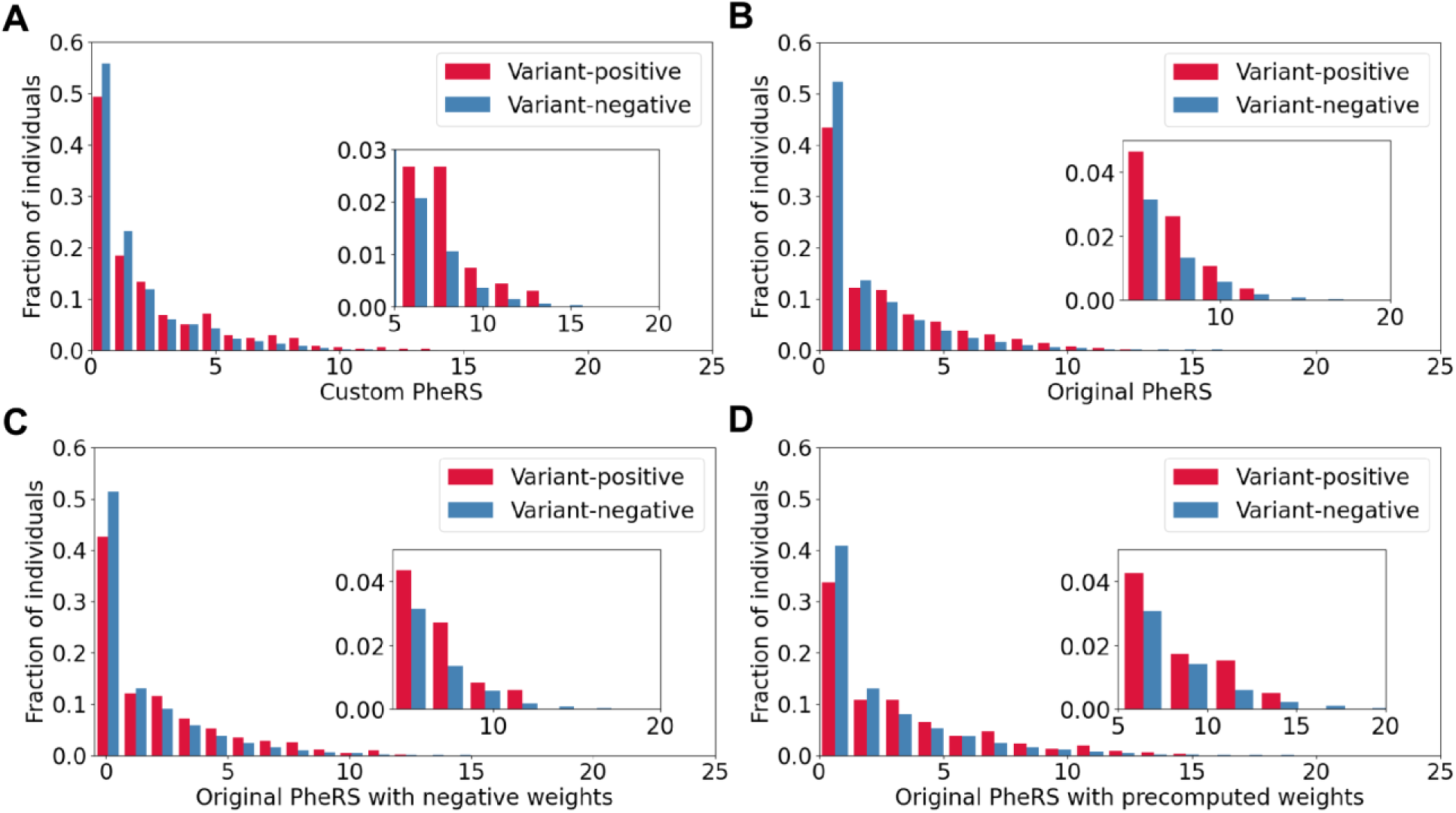
Distribution of PheRS among ATTRv V142I variant-positive and variant-negative individuals for the: **(A)** custom PheRS approach, **(B)** original PheRS approach, **(C)** original PheRS approach with negative weights, and **(D)** original PheRS approach with precomputed weights. For each plot, the inset shows the distribution in the high PheRS range ([5, 20]).

**Table 2.** Comparison of phenotypic risk score (PheRS) performance on the ATTRv cohort (cases: V142I+ individuals; controls: V142I-individuals). The value of *k* for precision-at-*k* was set to 100, i.e., 100 top-scoring individuals in terms of PheRS were considered.

| Method | Wilcoxon rank sum test P-value | AUC | Precision-at-*k* |
| --- | --- | --- | --- |
| Custom PheRS | $4.49 \times 10^{-4}$ | 0.552 | 0.050 |
| Original PheRS | $1.83 \times 10^{-6}$ | 0.571 | 0.000 |
| Original PheRS with negative weights | $1.73 \times 10^{-6}$ | 0.571 | 0.010 |
| Original PheRS with precomputed weights | $1.96 \times 10^{-6}$ | 0.571 | 0.010 |

In practice, an important clinical use for PheRS in ATTRv due to V142I would be the prioritization of individuals who are putative variant carriers and would benefit from genetic testing. To this end, we used the precision-at-*k* metric to quantify the effectiveness of different PheRS approaches in correctly identifying case individuals from among the high-scoring ones, without regard to low-scoring individuals. On this practically relevant metric, clear differences between methods emerged, with the custom PheRS approach performing the best when 100 individuals with the highest PheRS were considered (Table 2).

### Additional data-driven optimization modestly impacted PheRS performance

We further sought to improve and refine our custom PheRS approach through two typical data-driven methods. First, to identify the value of *k* at which predictive performance and utility are balanced for practical applications, we calculated precision for different *k* values, ranging from 10 to the size of the full cohort with different increment sizes. The custom PheRS approach not only outperformed the allele frequency of V142I in the cohort (which serves as a baseline), but also showed a 1.5- to 5-fold improvement over the original PheRS approach (Figure 2A). Notably, the differences between the custom and original approaches were the highest for smaller values of *k*, for which the original approach and its variants identified no variant-positive individuals. However, at small values of *k*, the precision value estimates are less stable as *k* tends to grow more rapidly than the number of true cases identified. At larger values of *k*, e.g., beyond 500 individuals, practical utility diminishes, as it may not be feasible to follow up on such a large number of individuals to identify the small number of variant-positive ones. While minor differences in trends were observed across the different algorithmic variations of the original PheRS approach, they converged to similar precision values as the custom approach once *k* reached around 1000 individuals. Taken together, we concluded that *k* values ranging from 100 to 200 represented the interval with the most practical utility and reliable improvements in predictive performance.

**Figure 2.**
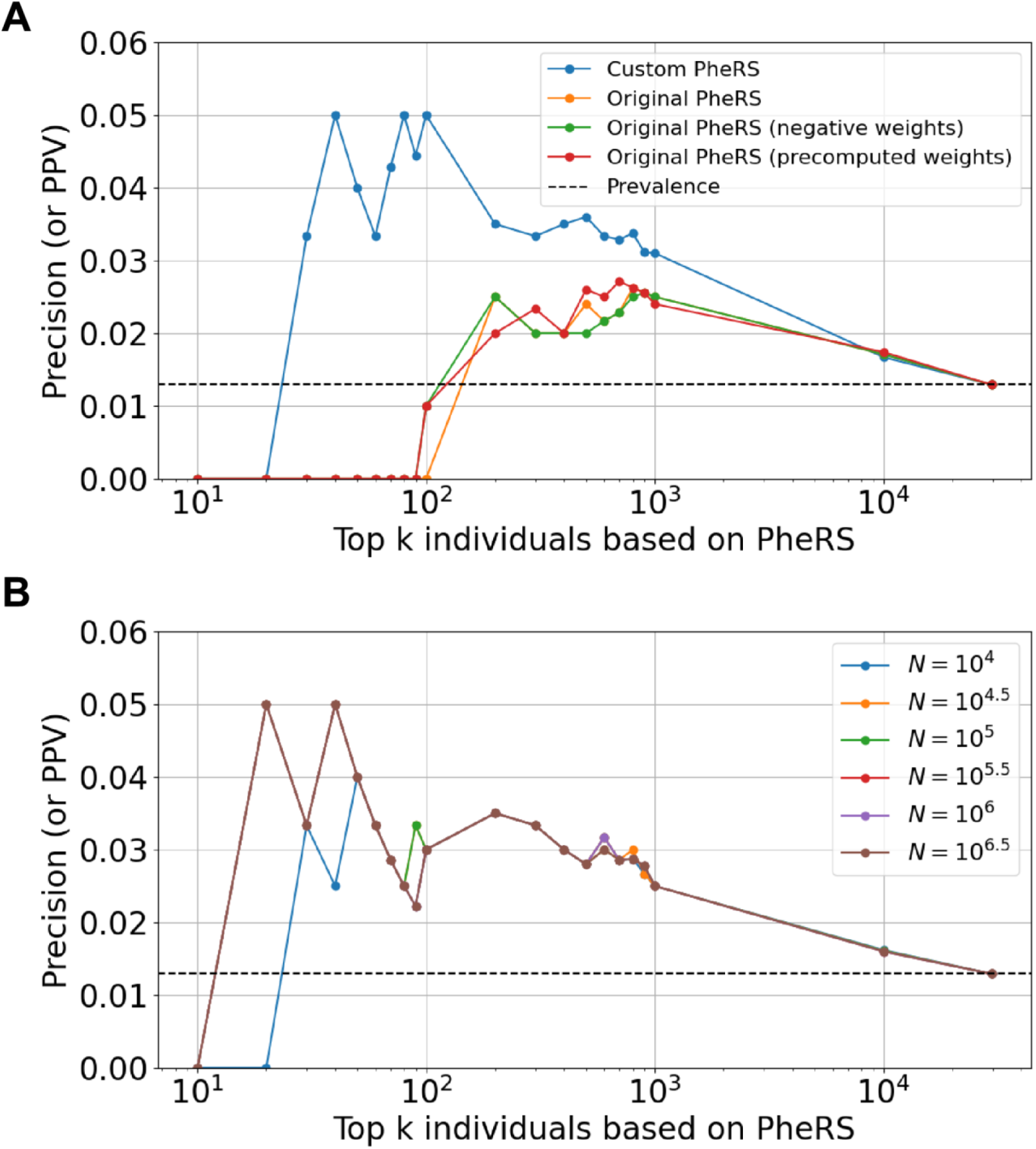
Refinement and optimization of PheRS through the assessment of different: **(A)** *k* values, and **(B)** training cohort sizes, i.e, the number of individuals used to compute the PheRS weights. For visualization purposes, the values of *k* were varied in increments of 10 in the interval [10, 100], increments of 100 in the interval [100, 1000] and then on the log scale (base 10) beyond that until the full cohort size was reached. The dashed black line corresponds to the prevalence of cases in the cohort (0.013) and will correspond to the precision when *k* = the size of the full cohort.

Next, we evaluated the impact of training cohort size on the performance of custom PheRS. To this end, we expanded the training cohort to include the entirety of the MSDW. We systematically varied the number of individuals used to compute PheRS weights, starting from a size similar in magnitude to that of our original (the Bio*Me*) training cohort to the entirety of MSDW. There was little to no difference in performance when the training cohort size was changed, with trends over *k* values remaining the same (Figure 2B). This is in agreement with trends observed for the original PheRS approach that uses weights precomputed from the BioVU biobank (Table 2, Figure 1D and Figure 2A).

### Specific knowledge about disease can improve PheRS performance and application

Given the age-based penetrance (symptoms typically emerge in the sixth decade of life^14^ or later) and population skew of V142I (this variant is found in 3-4% of African American individuals^16^), we assessed the impact of stratifying training and test cohorts by age and/or race/ethnicity on predictive performance. For age, we considered the subset of individuals who were at least 60 years old at the time of the last diagnosis in their EHR. For race/ethnicity, we considered the subset of individuals who were recorded as being African American in their EHRs. We trained and tested on all possible combinations of sub-cohorts and calculated precision-at-*k* values at *k* = 100 and *k* = 200. Another defining feature of V142I is its strong association with HF^13,15^, a major and early contributor of morbidity and mortality in this disease. We therefore further broke this analysis down in terms of the occurrence of a HF diagnosis in a patient’s history. Assuming that the ATTRv PheRS would be most useful in identifying patients who have been hospitalized for HF without any specific diagnostic testing for ATTRv, we only included individuals with at least one occurrence of a HF diagnosis in their EHRs. To avoid circularity, we recomputed the PheRS without the HF feature, effectively also testing the utility of PheRS for the identification of V142I cases before the occurrence of HF. We observed only minor differences in precision-at-*k* values between training on the full and various sub-cohorts. However, performance modestly improved when PheRS was applied to older African American individuals (Figure S1).

Since these observations were dependent on the choice of *k*, we assessed performance at multiple values of *k* within the optimal range as established above. We restricted the training and test cohorts to those with both age- and race/ethnicity-based constraints, with two baseline models as comparators: (1) a PheRS model trained on the full cohort, and (2) a model that randomly assigned case/control status based on prevalence in the test cohort. We observed modest differences between the model trained on the age- and race/ethnicity-specific cohort and that trained on the full cohort, with a slight advantage to the former when applied to an age- and race/ethnicity-specific test cohort (Figure 3A). Both models outperformed random guessing. These trends were more apparent when these sub-cohorts were further restricted to individuals with HF, with both models outperforming random guessing (Figure 3B). Stable peak precision-at-*k* values were observed at lower values of *k* when training on age- and race/ethnicity-specific cohorts than when training on the full cohort. While a higher precision-at-*k* value was reached relative to non-HF patients, it is not clear if this was a consequence of a higher prevalence of cases among patients with HF.

**Figure 3.**
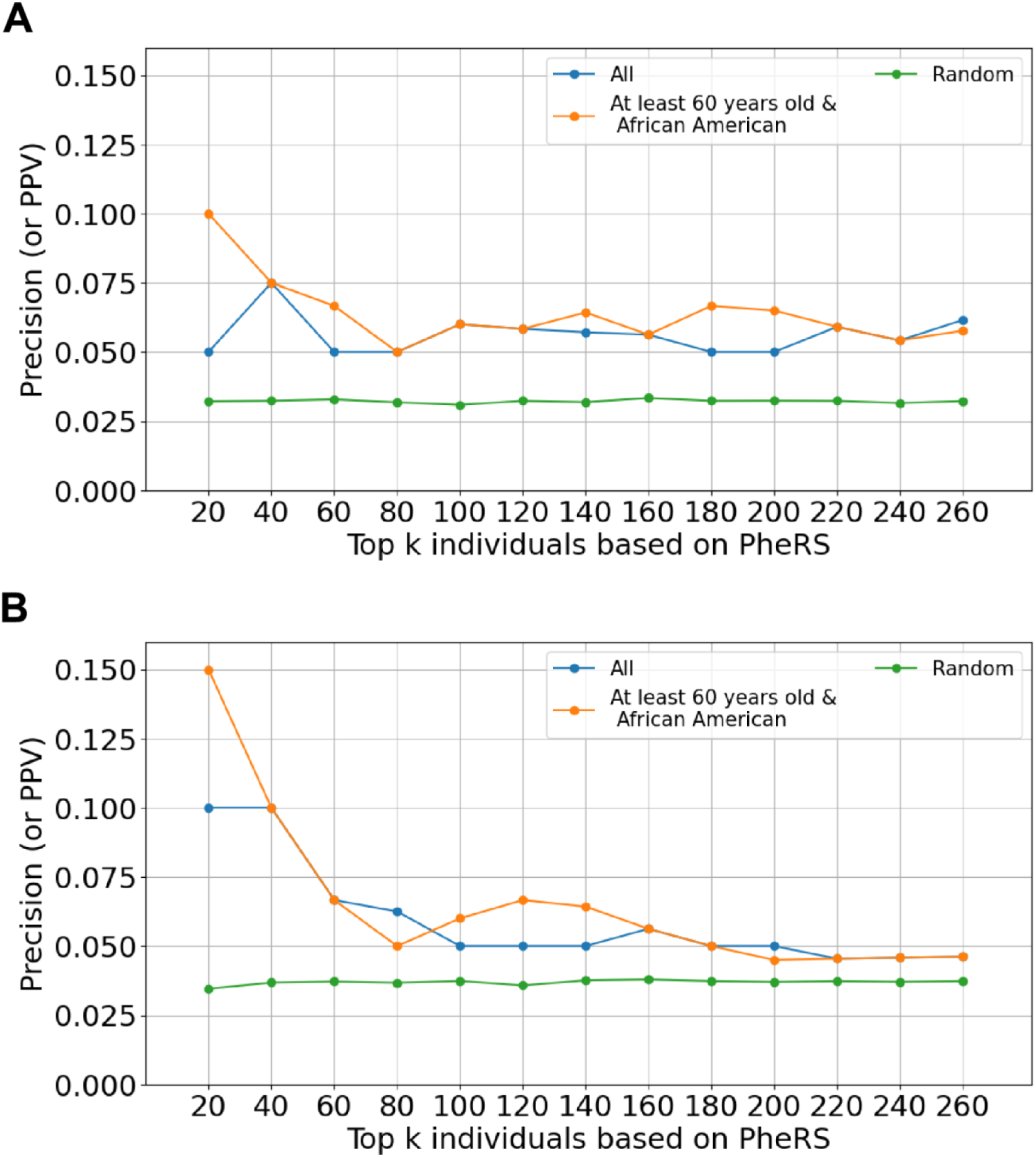
Precision-at-*k* values for PheRS after incorporating ATTRv-specific epidemiologic information into training and test cohort construction. In **(A),** the test/application cohort consisted of African American individuals who were at least 60 years old, and in **(B)** the test/application cohort consisted of African American individuals who were at least 60 years old and had at least one occurrence of a HF-related diagnosis code. In (B), the heart failure phenotype was excluded from PheRS calculation. Each curve corresponds to the demographic characteristics selected for in the training set. The random sampling approach assigns case-control status simply based on the prevalence of cases in the cohort. The most practical range of *k* values for clinical applications is shown here.

### Comparing and contrasting across other cardiovascular diseases

Finally, to test the generalizability of our observations with ATTRv, we expanded our comparison of custom PheRS approaches with standard PheRS to two more Mendelian diseases affecting the cardiovascular system (Tables S2 and S3). We identified individuals within the Bio*Me* biobank with putatively disease-causing variants for Marfan syndrome and Long QT syndrome (Table 1). As with ATTRv, individuals harboring putatively disease-causing variants were considered to be cases and those without such variants were considered to be controls.

As with ATTRv, the custom PheRS approach yielded higher precision values at lower values of *k*, outperforming other PheRS approaches (Figure 4). However, all methods performed similarly once *k* reached a large enough value. This value varied across diseases, with Long QT syndrome assessments exhibiting unusual trends in terms of the optimal range of *k* values. This results from the small sample sizes but may also be related to the penetrance of variants with respect to the phenotypes included in our PheRS. In the case of Marfan syndrome, since there are ICD codes for the actual disorder, we were able to test the penetrance hypothesis by constructing a PheRS calculator for the diagnosis code rather than *FBN1* variant presence or absence. Indeed, PheRS performance improved when constructed to distinguish diagnosed cases from undiagnosed controls (Figure S2).

**Figure 4.**
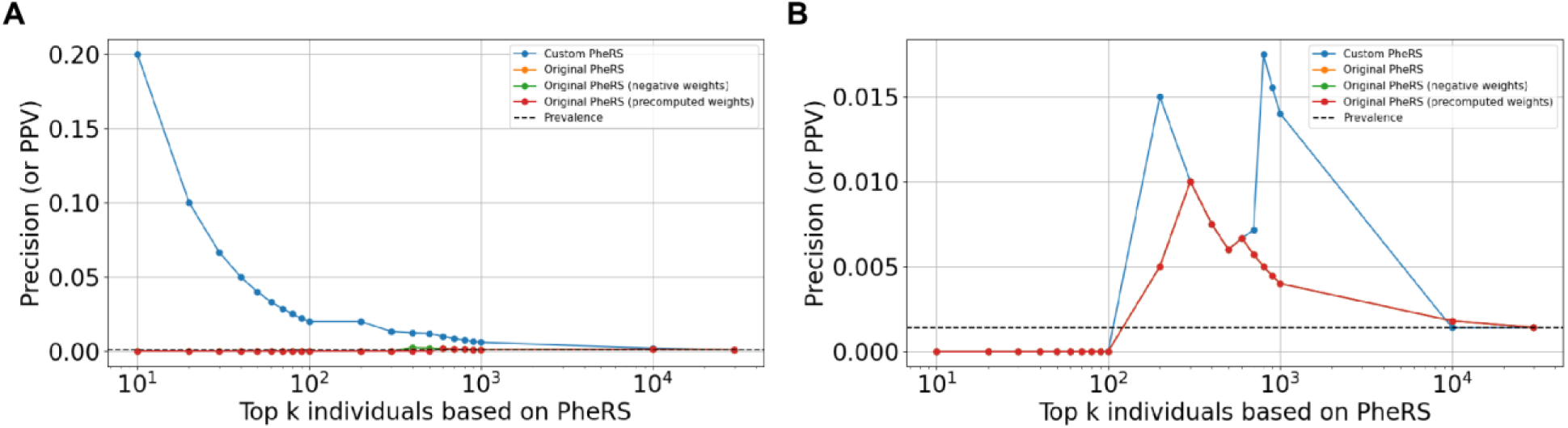
Precision-at-*k* values for the different PheRS approaches on cohorts for: **(A)** Marfan syndrome and **(B)** Long QT syndrome. The values of *k* were chosen in a manner similar to that in Figure 2A. The dashed black line corresponds to the prevalence of cases in each cohort and will correspond to the precision when *k* = the size of the full cohort.

## DISCUSSION

Monogenic cardiovascular disorders contribute to risk for HF, stroke, aneurysm, arrhythmia and sudden death and encompass a set of conditions that collectively carry high rates of morbidity and mortality. In large health systems and population databases linked to genomic sequencing data, ∼1:100 individuals harbor a putatively disease-causing variant associated with one of these disorders.^25^ A major limitation to delivering care that can mitigate associated risks of these conditions is the vast underutilization of genetic testing. For example, contemporary claims data have shown that genetic testing was provided to only 1-2% of patients with cardiomyopathies and inherited arrhythmias.^26^ PheRS is a promising tool that can potentially address the problems of underrecognition of these high risk disorders.

For this study, we considered traditional and emerging methods of measuring PheRS performance. In the literature, it is common to demonstrate that a proposed risk score is able to separate cases from controls by achieving statistically significant differences between the score distributions for these two groups.^11,22^ However, in the case of rare Mendelian disorders, this approach is likely to result in a type I error owing to the high case-control imbalance.^27^ This was indeed observed in this study, with a qualitative inspection of PheRS distributions and AUC comparisons suggesting that the significant separation between cases and controls was likely a false positive. On the other hand, AUC as a metric may be too stringent as it considers predictive performance on both ends of the PheRS distribution, which may not be necessary for the prioritization of patients for genetic testing. We envision PheRS as a tool that, when coupled with decision-support prompts in an EHR, can flag patients at risk for monogenic diseases who would benefit from genetic testing. A viable PheRS must therefore accurately identify individuals with relevant genotypes and minimize recommendations for genetic testing in genotype-negative individuals, i.e., minimize false positives and exhibit high specificity. However, due to the rarity of monogenic conditions, the number of false positives will almost always be low relative to the large number of genotype-negative individuals, ensuring relatively high specificity. The central question in this context then becomes: given a high PheRS for a patient, what is the probability that they harbor a likely pathogenic or pathogenic variant? Precision or positive predictive value at an appropriate PheRS threshold in terms of the estimated number of prioritized (top *k*-scoring) individuals, better addresses this particular question than sensitivity or specificity. Yet, since the prior probability of a random individual having a monogenic condition is low, precision-at-*k* values may still appear to be relatively small. This is especially true for the common V142I variant because of its incomplete penetrance, which practically makes the number of V142I+ individuals with ATTRv low. However, our precision-at-*k* estimates improved over the baseline prevalence, demonstrating that PheRS can indeed provide added clinical value without further enrichment of cases in the application population.

Standard PheRS features are derived from phecodes, which typically map to phenotype descriptions in the Online Mendelian Inheritance in Man (OMIM) database and may be broad and ambiguous. For ATTRv, the most heavily weighted phecodes in the standard PheRS R library included “nonspecific abnormal findings in cerebrospinal fluid,” “spinocerebellar disease,” “nystagmus and other irregular eye movements” and “abnormal reflex”, features that are not known to be associated with V142I-related disease. Furthermore, embedded in the phecodes for the standard PheRS were specific ICD-9 and ICD-10 codes for diagnoses that conflict with ATTRv, including wild-type ATTR and light chain amyloidosis (AL); co-occurance of these conditions with ATTRv is thought to be exceedingly rare and, therefore, their inclusion may contaminate PheRS calculations. Utilizing expert knowledge of V142I-associated ATTRv, we customized PheRS feature selection, which improved performance.

Phenotype-driven risk prediction scores have previously been tested for the purpose of prioritizing patients for genetic testing, but only for conditions that manifest with rare features (i.e., rare pediatric genetic diagnoses, chromosomal abnormalities).^28,29^ We sought to develop and refine a PheRS for V142I-associated ATTRv, a monogenic condition with manifestations that are relatively common in the general adult population, including HF, atrial fibrillation, carpal tunnel syndrome and neuropathy. Unlike some of the prior conditions in which PheRS have been applied, V142I-associated ATTRv is also characterized by incomplete and age-dependent penetrance. Although this poses additional challenges, we capitalized on highly specific ATTRv information to improve the performance of our approach. For example, since V142I is found in up to 4% of individuals with African ancestry^12^ and is extremely rare in non-African populations (0.0026% in non-Finnish Europeans in gnomAD), we tested the PheRS in this specific population. Furthermore, we used the knowledge that penetrance of V142I-related disease typically emerges after the sixth decade of life^14,19^ to focus our PheRS testing in an older adult population. Training and applying the model on an age- and race/ethnicity-specific cohort improved its performance and we saw even further improvements when we restricted the training and application to patients with HF. Therefore, a health systems-based implementation of PheRS targeting older, African American patients with HF may improve detection of V142I-associated ATTRv.

The practical performance of any PheRS, however, will depend on its implementation. Although PheRS represent promising new tools for recognizing undiagnosed diseases, several factors should be considered in their development and application across health systems, likely impacting their broad translation. First, models will be limited by the numbers of cases within a health system. We found that our PheRS for Marfan and Long QT syndromes exhibited less stable precision-at-*k* values than V142I ATTRv and lower performance than the standard PheRS for these disorders when considering other metrics (Figure 4; Table S5). This may be due to the smaller number of genotype-positive individuals for these conditions. Next, we found that a large number of cases across all three cohorts did not have any of the diagnosis codes included in the PheRS. This could be due to low penetrance, sampling bias or data quality issues such as information loss or corruption during transformation of EHR data. Third, PheRS for conditions that are rarer and have higher penetrance such as Marfan and Long QT syndromes require different choices of *k* values or PheRS thresholds (optimal *k* for Marfan syndrome was found to be 100 to 500 and for Long QT syndrome was found to be 800 to 1000). These are expected to yield different precision values relative to ATTRv, i.e., success of PheRS can be perceived differently in different contexts. Finally, the development and assessment of models often needs to be aligned with implementation objectives, limiting a single generalizable approach across health systems and applications. For example, while the original PheRS approach relied on entire patient populations to calculate model weights, we found that training cohort size had little to no influence on predictive performance. In fact, there was a slight drop in overall performance at a given *k* value in the MSDW training versus the Bio*Me* training cohorts. The Bio*Me* cohort tends to skew older, and is likely to be more representative of the population on which the ATTRv PheRS would be applied.

In summary, PheRS have the potential to usher in precision cardiology by identifying individuals with high-risk Mendelian disease genotypes in health systems, but benefit from customization considering well-described genotype-specific clinical and demographic features. Importantly, since the PheRS approach uses only ICD codes and demographic data, it is more feasible to implement across health systems, particularly low-resourced clinics where many undiagnosed patients receive care, compared with algorithms that use more complex data elements. As >1.5 million Americans harbor the *TTR* V142I variant and >85% of symptomatic carriers remain undiagnosed^14^, the ATTRv PheRS may be a useful tool to improve recognition of risk, shortening time to diagnosis and treatment.

## Data Availability

The datasets analyzed for the study are not publicly available. The Genomic and EHR data cannot be redistributed to researchers other than those approved through the Icahn School of Medicine at Mount Sinai (ISSMS) Institutional Review Board. We have therefore given detailed description in the manuscript. The code will be made available on GitHub at the time of publication.

## ACKNOWLEDGEMENTS

We thank Ms. Briana Christian for help with knowledge-based mapping of ATTRv ICD codes for PheRS. We also thank Dr. Mariya Shadrina for help with variant identification and curation.

